# Changes in thoracic Cobb angle dynamics following Nuss procedure and bar removal in adult patients with pectus excavatum

**DOI:** 10.1101/2025.08.05.25333059

**Authors:** Nay Htut, Der-En Keong, I-Shiang Tzeng, Yeung-Leung Cheng

**Affiliations:** Division of Thoracic Surgery, Department of Surgery, Taipei Tzu Chi Hospital, Buddhist Tzu Chi Medical Foundation, New Taipei City, Taiwan; Department of Research, Taipei Tzu Chi Hospital, Buddhist Tzu Chi Medical Foundation, New Taipei City, Taiwan; Department of Statistics, National Taipei University, New Taipei City, Taiwan; School of Medicine, Tzu Chi University, Hualien, Taiwan

**Author notes:** **Corresponding author:** (Y-L C).

## Abstract

**Background:** Pectus excavatum is associated with spinal abnormalities. The Nuss procedure is the standard surgical correction approach, though its effect on spinal alignment, especially the thoracic Cobb angle, remains unclear. This study examined changes in thoracic Cobb angle dynamics in 109 adult patients with pectus excavatum.

**Methods:** Patients with pectus excavatum (109; 86 males and 23 females; mean age: 23.9 ± 5.29 years) who underwent the Nuss procedure and subsequent bar removal were retrospectively analyzed. Demographic and clinical data were collected, including serial thoracic Cobb angle measurements from posteroanterior chest radiographs preoperatively, 1 month, 3–6 months, and 1 year postoperatively, 1 day pre-bar removal, and 1 week post-bar removal. Subgroup analyses were stratified by sex, Haller index (≥ or <4), bar number (1, 2, or 3) and position (oblique vs. horizontal), bar flipping within 3 months, and preoperative Cobb angle (< or ≥10°).

**Results:** Mean thoracic Cobb angle significantly increased 1 month postoperatively (preoperative: 5.35±4.80° vs. 1 month postoperative: 5.75±5.00°, P=0.004), but significantly decreased 1 day before bar removal (preoperative: 5.35±4.80° vs. 1 day pre-bar removal: 4.90±4.74°, P=0.006) and 1 week after (preoperative: 5.35±4.80° vs. 1 week post-bar removal: 4.26±4.72°, P<0.001). Cobb angle significantly improved after bar removal.

**Conclusions:** In adult patients with pectus excavatum, the Nuss procedure induced a transient increase in thoracic Cobb angle 1 month postoperatively, followed by a significant reduction after bar removal. Early bar flipping correlated with diminished Cobb angle correction.

## Introduction

Pectus excavatum (PE), or funnel chest, accounts for approximately 65–95% of chest wall anomalies; it is characterized by posterior displacement of the sternum and adjacent costal cartilages affecting up to eight in 1,000 births, with a 5:1 male predominance [1,2]. PE is not only a cosmetic concern but is also associated with cardiopulmonary compromise and significantly increased scoliosis prevalence [3]. Although the Nuss procedure effectively corrects the thoracic depression, its effect on spinal alignment remains controversial. Previous studies have reported conflicting outcomes regarding Cobb angle (CA) changes following the Nuss procedure in patients with PE, with reports of both reduction and progression. Most available evidence focuses on pediatric and adolescent populations [4–10], while data on adult patients remain scarce. This retrospective study examined the longitudinal thoracic CA dynamics in 109 adult patients with PE undergoing the Nuss procedure with subsequent bar removal (BR), evaluating both clinical (sex and Haller index) and surgical (bar number, position, and flipping) factors influencing spinal alignment.

## Materials and Methods

### Patient Cohort and Data Collection

We conducted a retrospective review of 109 consecutive patients with PE (86 males and 23 females; mean age: 23.91 ± 5.29 years) who underwent the Nuss procedure with subsequent BR at Taipei Tzu Chi Hospital between August 2014 and December 2020, with inclusion criteria of age ≥ 20 years at surgery, completed BR, and complete clinical and radiological data; patients with prior spinal surgery were excluded. Collected data encompassed demographic and anthropometric measures (sex, age, height, weight, body mass index, and preoperative Haller index) and surgical variables (number of bars [1–3], bar position [horizontal or oblique], and bar flipping within 3 months). Horizontal bar placement was defined as both ends positioned within the same intercostal space, whereas oblique bar placement required the ends to occupy different intercostal spaces. Bar flipping was defined as a slope angle (α) > 30° according to Fan et al.’s criterion [11].

This study was approved by the Institutional Review Board and Ethics Committee of Taipei Tzu-Chi Hospital, Taipei, Taiwan (no. 14-IRB012). The need for informed consent was waived due to the retrospective nature of the study.

### Thoracic CA Measurement

CAs were measured manually on standing posterior-anterior chest radiographs of thoracic vertebrae T6–T12 (Figs 1 and 2) at six timepoints: preoperatively, 1 month, 3–6 months, and 1 year postoperatively, 1 day pre-BR, and 1 week post-BR. Two blinded independent observers performed all measurements, and interobserver discrepancies were resolved through averaging and consensus review.

**Fig 1.**
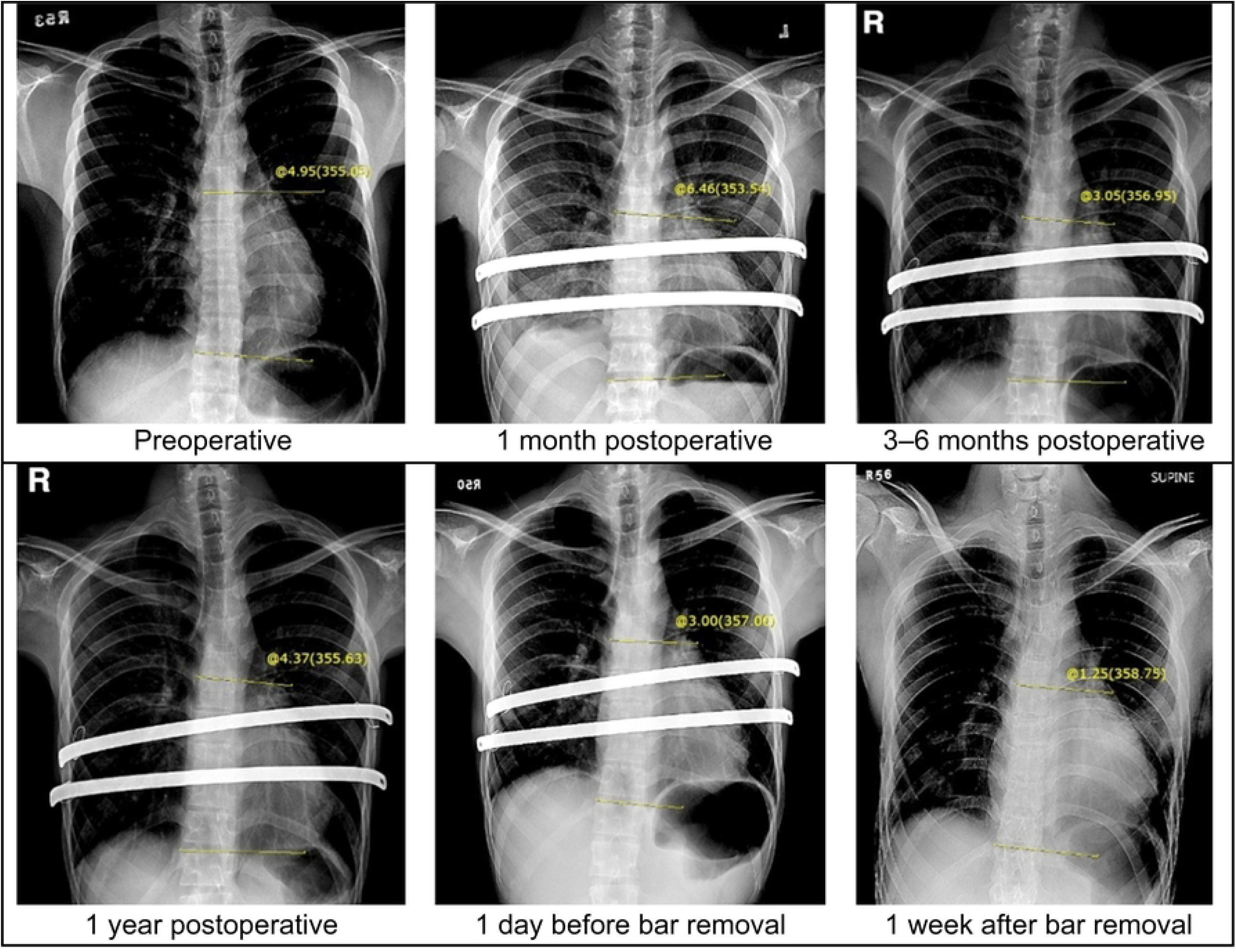
Thoracic Cobb angles were measured on posteroanterior chest radiographs at six defined timepoints in patients with preoperative thoracic Cobb angles < 10°

**Fig 2.**
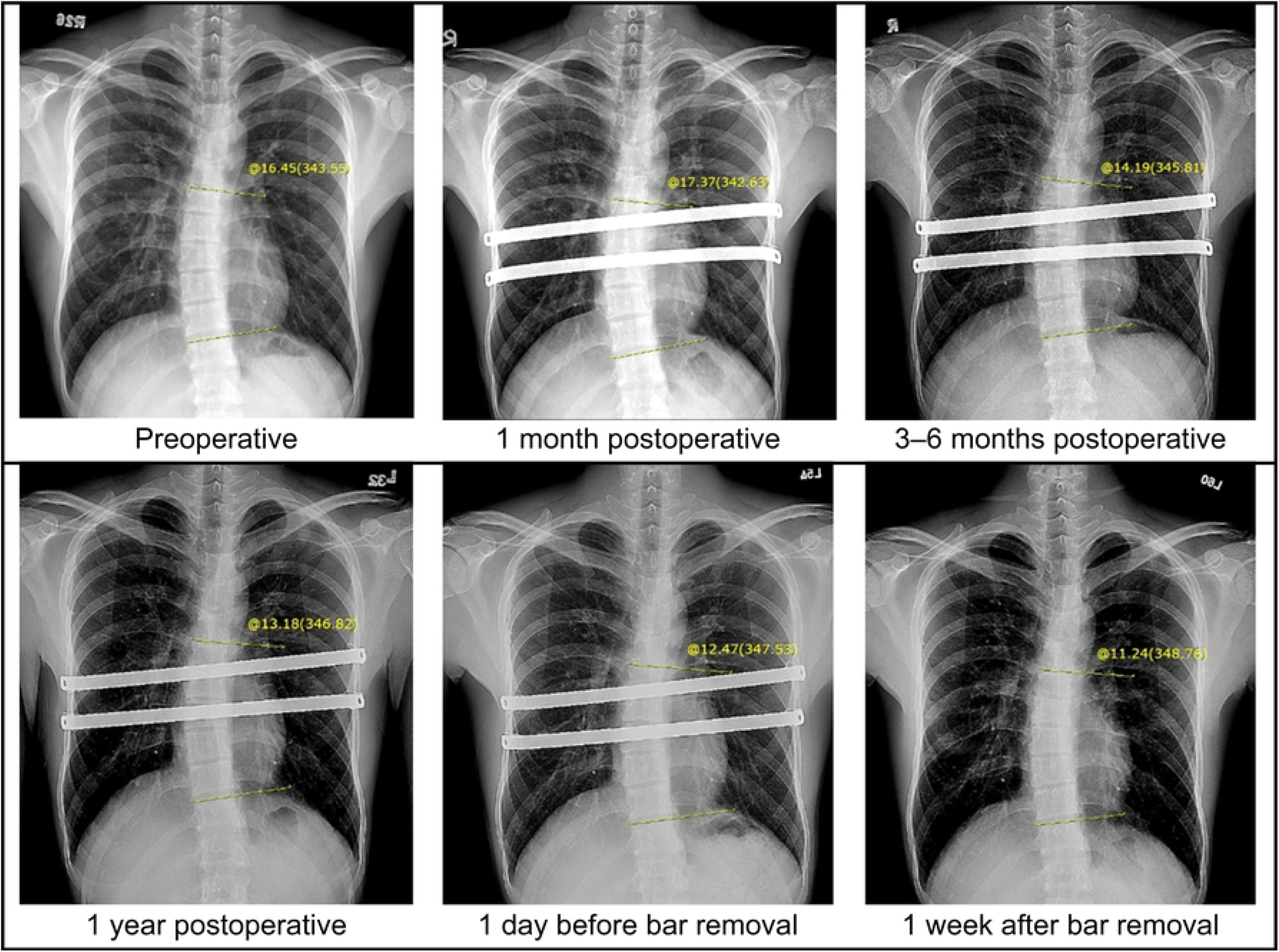
Thoracic Cobb angles were measured on posteroanterior chest radiographs at six defined timepoints in patients with preoperative thoracic Cobb angles > 10°

### Statistical Analysis

Continuous variables are presented as mean ± standard deviation and range. Categorical variables are presented as counts and percentages. Paired t-tests with Bonferroni adjustments were used for pairwise comparisons. Subgroup analyses were conducted using independent t-tests or one-way analysis of variance (ANOVA) as appropriate. Statistical significance was set at *p* < 0.05. To ensure confidentiality, all research data were delinked prior to analysis. Primary statistical analyses were performed using an artificial intelligence analytical platform (Perplexity pro version), and independent validation was conducted using SPSS (version 24; IBM Corp.).

## Results

Patient demographics and clinical characteristics are presented in Table 1. In 109 patients, the thoracic CA increased significantly at 1 month postoperatively compared to preoperative measurements (preoperative CA: 5.35 ± 4.80 vs. 1 month postoperative CA: 5.75 ± 5.00, P = 0.004). However, it decreased significantly 1 day pre-BR (preoperative CA: 5.35 ± 4.80 vs. 1 day pre-BR: 4.90 ± 4.74, P = 0.006) and 1 week post-BR (preoperative CA: 5.35 ± 4.80 vs. 1 week post-BR: 4.26 ± 4.72, P < 0.001) (Table 2).

**Table 1.** Patient demographics and clinical characteristics.

| Characteristic | Mean ± SD or n (%) |
| --- | --- |
| Number of patients | 109 |
| Age at correction (years) | $23.91 \pm 5.29$ |
| Preoperative CA ($^{\circ}$) | $5.35 \pm 4.8$ |
| Range | 0.39–31.4 |
| Median | 3.46 |
| Sex |  |
| Male | 86 (78.90%) |
| Female | 23 (21.10%) |
| Haller index | $4.13 \pm 1.85$ |
| Number of bars |  |
| 1 | 14 (12.84%) |
| 2 | 75 (68.81%) |
| 3 | 20 (18.35%) |
| Bar position |  |
| Horizontal | 58 (53.21%) |
| Oblique | 51 (46.79%) |
| Bar flipping within 3 months |  |
| Yes | 13 (11.93%) |
| No | 96 (88.07%) |
| Preoperative CA | 5.35 ± 4.80 |
| Patient height (cm) | 173.39 ± 7.35 |
| Patient weight (kg) | 60.05 ± 9.17 |
| BMI | 19.92 ± 2.39 |

BMI, body mass index; SD, standard deviation; CA, Cobb angle

**Table 2.** Thoracic CA changes over time.

| Timepoint | Mean CA (°) ± SD | Δ vs. Preoperative (°)<br>(%) | p-value |
| --- | --- | --- | --- |
| Preoperative | 5.35 ± 4.80 | - | - |
| 1 month postoperative | 5.75 ± 5.00 | +0.41 (+7.63%) | 0.004* |
| 3–6 months postoperative | 5.27 ± 5.21 | -0.11 (-2.04%) | 0.464 |
| 1 year postoperative | 5.17 ± 5.04 | -0.25 (-4.59%) | 0.155 |
| 1 day pre-BR | 4.90 ± 4.74 | -0.44 (-8.28%) | 0.006* |
| 1 week post-BR | 4.26 ± 4.72 | -1.06 (19.87%) | < 0.001* |

Comparison of the preoperative CA with that at five subsequent timepoints.
BR, bar removal; Cobb angle, CA; Δ, change; SD, standard deviation.

### Subgroup analysis

Sex-stratified analyses revealed that males (n = 86) exhibited significant CA increases at 1 month postoperatively (P = 0.008) but decreases at 1 day pre-BR (P = 0.008) and 1 week post-BR (P < 0.001), whereas females (n = 23) showed decreases only at 1 week post-BR (P < 0.001). Those with a Haller index ≥ 4 (n = 43) had significant CA decreases both 1 day pre-BR (P = 0.003) and 1 week post-BR (P < 0.001), while those with an index < 4 (n = 66) had increases at 1 month postoperatively (P = 0.01) and decreases at 1 week post-BR (P < 0.001). Patients with one (n = 14) and three bars (n = 20) exhibited significant decreases in CA at 1 week post-BR (P = 0.001 and 0.003, respectively). In patients with two bars (n = 75), CA increased at 1 month postoperatively (P = 0.001) but decreased at 1 day pre-BR (P = 0.01) and 1 week post-BR (P < 0.001). Patients with horizontal bars (n = 58) demonstrated a significant increase in CA at 1 month postoperatively (P < 0.001) but decreased CA at 1 week post-BR (P < 0.001), while those with oblique bars (n = 51) had decreased CA at 1 day pre-BR (P = 0.023) and 1 week post-BR (P < 0.001). Patients who experienced bar flipping within 3 months postoperatively (n = 13) showed CA reduction only at 1 week post-BR (P = 0.02); in contrast, patients without bar flipping (n = 96) exhibited an increase in CA 1 month postoperatively (P = 0.008), followed by a decrease 1 day pre-BR (P = 0.004) and 1 week post-BR (P < 0.001). Patients with preoperative CA stratification ≥ 10° (n = 17) had CA decrease both 1 day pre-BR (P = 0.017) and 1 week post-BR (P = 0.022), while those with preoperative CA < 10° (n = 92) had CA increase at 1 month postoperatively (P = 0.002) but decrease at 1 week post-BR (P < 0.001) (see Supplementary file, Tables S1–S6).

## Discussion

While the Nuss procedure transiently exacerbated CAs (likely due to early postoperative pain and discomfort caused by bar insertion and thoracic expansion forces), BR restored near-baseline alignment with a reduction in thoracic CA. Females showed only a significant reduction at 1 week post-BR, possibly because of an inadequate number of cases. All subgroups demonstrated significant CA reduction 1 week post-BR. However, patients who experienced bar flipping within the first 3 postoperative months had a less favorable reduction in CA. Additional findings revealed that female patients had significantly higher preoperative CAs than those in males (7.58 ± 6.42 vs. 4.75 ± 4.11, respectively; P = 0.022), and patients with a Haller index ≥ 4 had greater preoperative CAs than those with an index < 4 (6.65 ± 5.74 vs. 4.49 ± 3.89, P = 0.034).

Ghionzoli, et al. [4] reported a mean thoracic CA reduction of 1.5° (from 10.9° to 9.4°) following complete treatment (Nuss procedure with subsequent BR) in patients with PE with adolescent idiopathic scoliosis. Our adult cohort demonstrated a comparable correction magnitude (mean reduction of 1.06°), representing a 19.87% decrease in CA. Park, et al. [5] observed a decrease in CA after corrective surgery (Nuss procedure followed by BR) in the early correction group (age at surgery < 10 years) but an increase in the late correction group (age at surgery ≥ 10 years). These patients were primarily children or adolescents, most of whom had either a spinal curve (0–10°) or mild scoliosis (10–20°) [12]. In contrast, our study found that all adult patients exhibited a significant reduction in CA after BR, including those with a CA ≥ 10°. Chung, et al. (2017) reported that preoperative CA > 15° was associated with postoperative CA progression, whereas preoperative CA < 15° was associated with postoperative CA reduction [6]. In the current study, most patients had a preoperative CA < 15° and demonstrated a reduction in CA after BR.

Niedbala, et al. [13] reported two cases of acquired scoliosis following the Nuss procedure, with CAs of 14° and 16°, which were attributed to potential acute asymmetric pressure resulting from Nuss repair [13]. Meng, et al. [7] reported a case of acquired scoliosis in a 14-year-old male that developed within 1 month after the Nuss procedure, with rapid progression to a severe CA of 54°. This pattern of early postoperative CA progression (within 1 month) is consistent with our study findings. Floccari, et al. [8] described two cases of PE in patients whose severe scoliosis progressed to a surgical-range deformity (CA of 49–75° and 39–65°, respectively) within 3 months postoperatively following the Nuss procedure. İşcan, et al. [9] reported divergent outcomes regarding CA progression following complete PE correction treatment. Although CA increased significantly in all patients overall, particularly in males and children, no significant increase was observed in adults or females.

The postoperative progression of CA after the Nuss procedure was associated with moderate to severe scoliosis (CA ≥ 20°) [6,8,12]. Adult patients with a spinal curve and mild scoliosis (CA < 20°) [12] receive benefit from CA reduction after complete PE correction surgery. The limitations of this study were its single-center retrospective design and limited number of study cases. Large-scale, multicenter studies are warranted to better evaluate the impact of the Nuss procedure on thoracic CA in adult patients with pectus excavatum.

## Conclusion

The thoracic CA in adult patients showed a transient increase 1 month after the Nuss procedure, but a reduction was observed following BR compared with preoperative values. Subgroup analyses consistently demonstrated a significant decrease in CA after BR. These findings indicate that adult patients with mild or less severe scoliosis with PE who undergo the Nuss procedure and subsequent BR may experience improvement in thoracic spinal alignment, potentially reducing the risk of scoliosis progression.

## Data Availability

All relevant data are within the manuscript and its Supporting Information files.

## Acknowledgments

Not Applicable.

## Supporting information

**Table S1. Dynamic changes in thoracic CA by sex**

**Table S2. Dynamic changes in thoracic CA by HI**

**Table S3. Dynamic changes in thoracic CA by number of bars**

**Table S4. Dynamic changes in thoracic CA by bar position**

**Table S5. Dynamic changes in thoracic CA by bar flipping within 3 months**

**Table S6. Dynamic changes in thoracic CA by preoperative CA ≥ and < 10°**

## Notes

### Competing Interest Statement

The authors have declared that no competing interests exist.

### Funding Statement

Yes

### Author Declarations

This study was approved by the Institutional Review Board and Ethics Committee of Taipei Tzu-Chi Hospital, Taipei, Taiwan (IRB No: 14-IRB012). The need for informed consent was waived due to the retrospective nature of the research.

